# Functional connectivity of white matter as a biomarker of cognitive decline in Alzheimer’s disease

**DOI:** 10.1101/2020.05.05.20091892

**Authors:** Yurui Gao, Anirban Sengupta, Muwei Li, Zhongliang Zu, Baxter P. Rogers, Adam W. Anderson, Zhaohua Ding, John C. Gore, for the Alzheimer’s Disease Neuroimaging Initiative

## Abstract

**Objective:** *In vivo* functional changes in white matter during the progression of Alzheimer’s disease (AD) have not been previously reported. Our objectives are to measure changes in white matter functional connectivity (FC) in an aging population undergoing cognitive decline as AD develops, to establish their relationship to neuropsychological scores of cognitive abilities, and to assess their performance as predictors of AD.

**Methods:** Analyses were conducted using resting state functional MRI (rsfMRI) and neuropsychological data from 383 ADNI participants, including 136 cognitive normal (CN) controls, 46 with significant memory concern, 83 with early mild cognitive impairment (MCI), 37 with MCI, 46 with late MCI, and 35 with AD dementia. We used novel analyses of whole brain rsfMRI data to derive FC metrics between white matter tracts and discrete cortical volumes, as well as FC metrics between different white matter tracts, and their relationship to 6 cognitive measures. We then implemented supervised machine learning on white matter FCs to classify the participants and evaluated the performance.

**Results:** Significant decreases were found in white matter FCs with prominent, specific, regional deficits appearing in late MCI and AD dementia patients relative to CN. These changes significantly correlated with behavioral measurements of impairments in cognition and memory. The sensitivity and specificity for distinguishing AD dementia and CN using white matter FCs were 0.83 and 0.81 respectively.

**Conclusions and Relevance:** The white matter FC decreased in late MCI and AD dementia patients compared to CN participants, and the white matter FC correlates with cognitive measures. White matter FC based classification shows promise for differentiating AD patients from CN. It is suggested that white matter FC may be a novel imaging biomarker of AD progression.

## Introduction

Alzheimer’s disease (AD) is the most common progressive neurodegenerative disorder, which begins at a pre-symptomatic stage before subjects exhibit increasingly severe cognitive impairments and ultimately, dementia [1, 2]. Histopathological evidence of degeneration during this progression has been observed in human brains in both gray matter (GM) and white matter (WM) [3, 4]. While there has been considerable emphasis on cortical changes, pathological alterations of WM post mortem have also been reported not only in late stages of AD (associated with loss of axons and oligodendrocytes [5] and concomitant with vascular abnormalities [6, 7]), but also in earlier, pre-clinical stages, probably related to amyloid toxicity [8]. Moreover, tissue shrinkage has been found to be even more prominent in WM than in GM in early stage disease [3]. Potentially, therefore, appropriate measures of changes within WM may be valuable biomarkers of neurodegeneration in AD. However, to date there have been no studies of functional changes in WM in AD, and there remain no imaging metrics from any modality that reliably reflect behavioral or cognitive changes in the progression towards AD.

Functional magnetic resonance imaging (fMRI) has been previously used to detect functional alterations arising in AD [9–11] but to date such studies have been almost exclusively focused on GM, with very limited exceptions [12, 13]. FMRI detects changes in MRI images caused by variations in blood volume and/or oxygenation (blood oxygenation level dependent (BOLD) effects) that in cortex correspond to changes in neural activity. Correlations across time between fluctuations in MRI signals from different cortical regions in a resting state are interpreted as indicators of functional connectivity (FC) between them. BOLD signals are expected to be much smaller in WM than in GM and are usually excluded from image analyses. However, recently we demonstrated that BOLD fluctuations in WM share common features with those from GM and they correlate significantly with BOLD signals from specific cortical regions to which they connect [14, 15]. Relationships between WM tracts and GM regions may be summarized by a functional correlation matrix (FCM) of their pair-wise correlations at rest, while different WM tracts can be inter-related using a similar approach.

In this study, we extended these new findings and analyses, originally described in reference [14, 16], to quantify changes in WM fMRI metrics during the progression to AD. We measured the differences in the FCMs for WM-GM correlations (FCM_WG_) and WM-WM (FCM_WW_) between a healthy group and each of five participant groups at different stages of cognitive impairments. We also subsequently evaluated the correlations between these FC metrics and behavioral measures of cognition and memory. Finally, we explored the use of machine learning to differentiate between the controls and patients at different AD stages to evaluate how well WM FC alone can classify subjects.

## Materials and Methods

Data used in this study were all obtained from the database of the Alzheimer’s Disease Neuroimaging Initiative (ADNI) (adni.loni.usc.edu).

### Participants

The participant selection criteria were that 1) baseline rsfMRI images were available in ADNI-2 or ADNI-3 (if the participant was available in both ADNI-2 and ADNI-3, then data in ADNI-3 were selected); 2) participants were aged 60-90 years; 3) multi-band rsfMRI acquisitions were excluded and 4) the data survived MRI processing.

The participants were grouped as cognitive normal (CN), significant memory concerns (SMC), early mild cognitive impairments (EMCI), mild cognitive impairments (MCI), late MCI (LMCI) and AD dementia (ADD). The full criteria for clinical classifications are described in the ADNI manual [17].

### MRI

3T rsfMRI and T1-weighted (T1w) data, acquired at multiple institutions with the same imaging protocol, were preprocessed using the toolbox DPARSFA [18]. First, the rsfMRI images were corrected for slice timing and head motion. Twenty-four motion parameters [19] and mean CSF signal were regressed out. The resulting rsfMRI data were filtered (passband=0.01-0.1Hz), coregistered to a common space [20], detrended, and then normalized voxel-wise into a time-course with zero mean and unit variance. Next, WM, GM, and cerebrospinal fluid (CSF) were segmented using the T1w images [21] and their tissue probability maps were normalized to the common space.

### Calculation of Functional Correlation Matrix (FCM)

The calculations of correlations for each participant were restricted to WM and GM regions of interest (ROIs, listed in **Table 1**) that were defined by the Eve atlas [22] (48 WM tracts, see **S1 Fig** in Supplement) and PickAtlas [23] (82 Brodmann areas) and were further constrained within masks generated by thresholding the WM and GM probability maps at 0.8. The preprocessed time-courses were averaged over the voxels within each ROI and for each pair of WM and GM ROIs they were then cross correlated, excluding any time points with large motions (framewise displacement [24] >0.5). The resulting 48×82 correlation coefficients formed an FCM of WM-GM pairs (FCM_WG_). Similarly, the mean time-courses for each pair of WM ROIs were cross correlated and the 48×48 correlation coefficients formed an FCM of WM-WM pairs (FCM_WW_). Meanwhile, we generated FCM of GM-GM pairs (FCM_GG_) to confirm the validity of our processing pipeline. The possible influences of age, gender, years of education and acquisition-site were regressed out from FCM using a generalized linear model.

**Table 1.** List of WM and GM ROIs

| White Matter (WM) ROIs | Gray Matter (GM) ROIs |
| --- | --- |
| CST: Corticospinal Tract | BA1: Primary Somatosensory Cortex 1 |
| ML: Medial Lemniscus | BA2: Primary Somatosensory Cortex 2 |
| ICP: Inferior Cerebellar Peduncle | BA3: Primary Somatosensory Cortex 3 |
| SCP: Superior Cerebellar Peduncle | BA4: Primary Motor Cortex |
| CP: Cerebral Peduncle | BA5: Somatosensory Association Cortex |
| ALIC: Anterior Limb of Internal Capsule | BA6: Premotor and Supplementary Motor |
| PLIC: Posterior Limb of Internal Capsule | BA7: Visuo-Motor Coordination |
| RLIC: Retrolenticular Limb of Internal Capsule | BA8: Frontal Eye Fields |
| ACR: Anterior Corona Radiata | BA9: Dorsolateral Prefrontal Cortex |
| SCR: Superior Corona Radiata | BA10: Anterior Prefrontal Cortex |
| PCR: Posterior Corona Radiata | BA11: Orbitofrontal Area |
| PTR: Posterior Thalamic Radiation (include OR) | BA13: Insular Cortex |
| SS: Sagittal Stratum (include inferior longitudinal fasciculus and fronto-occipital fasciculus) | BA17: Primary Visual Cortex (V1) |
| EC: External Capsule | BA18: Secondary Visual Cortex (V2) |
| CGG: Cingulum (Cingulate Gyrus) | BA19: Associative Visual Cortex (V3-5) |
| CGH: Cingulum (Hippocampus) | BA20: Inferior Temporal Gyrus |
| FXC: Fornix (Cres) | BA21: Middle Temporal Gyrus |
| SLF: Superior Longitudinal Fasciculus | BA22: Superior Temporal Gyrus |
| SFO: Superior Fronto-Occipital Fasciculus | BA23: Ventral Posterior Cingulate Cortex |
| UF: Uncinate Fasciculus | BA24: Ventral Anterior Cingulate Cortex |
| TAP: Tapetum | BA25: Subgenual Area |
| MCP: Middle Cerebellar Peduncle | BA26: Ectosplenial Portion of Retrosplenial Region |
| PCT: Pontine Crossing Tract | BA27: Piriform Cortex |
| GCC: Genu of Corpus Callosum | BA28: Ventral Entorhinal Cortex |
| BCC: Body of Corpus Callosum | BA29: Retrosplenial Cingulate Cortex |
| SCC: Splenium of Corpus Callosum | BA30: Part of Cingulate Cortex |
| FX: Fornix | BA32: Dorsal Anterior Cingulate Cortex |
|  | BA34: Dorsal Entorhinal Cortex |
|  | BA35: Perirhinal Cortex |
|  | BA36: Ectorhinal Area |
|  | BA37: Occipitotemporal Area (part of fusiform gyrus and interior temporal gyrus) |
|  | BA38: Temporopolar Area |
|  | BA39: Angular Gyrus |
|  | BA40: Supramarginal Gyrus |
|  | BA41: Auditory Cortex 1 |
|  | BA42: Auditory Cortex 2 |
|  | BA43: Primary Gustatory Cortex |
|  | BA44: Pars Opercularis |
|  | BA45: Pars Triangularis |
|  | BA46: Dorsolateral Prefrontal Cortex |
|  | BA47: Pars Orbitalis |

### Neuropsychological Testing

Neuropsychological scores included the scores of the Mini-Mental State Examination (MMSE), Clinical Dementia Rating (CDR) global, CDR sum of boxes (CDR-SOB), Global Deterioration Scale (GDS), functional assessment questionnaire (FAQ) total, Wechsler memory scale-logical memory II subscale (WMS-LMII), Alzheimer’s Disease Assessment Scale-Cognitive (ADAS-Cog) and Hachinski scale, which are the most commonly used for clinical assessment and AD studies.

### Statistical Analysis

The characteristics of the six subject groups were summarized, and the differences among groups were tested by one-way ANOVA or chi-squared test.

The FCMs within each clinical group were averaged to produce a mean matrix (mFCM). Differences in the mFCM values, and the effect sizes of these differences [25] between the CN group and every other group were calculated. Permutation tests (10,000 permutations) were conducted for each FCM element across all participants within any two comparison groups. The resulting *P*-values were corrected for multiple comparisons using a false discovery rate [26], denoted as P_fdr_. To estimate the overall connectivity of each WM tract, the FCM elements corresponding to each WM ROI were averaged. The mean and standard deviation of each WM-tract-wise FC across participants within each group were then calculated. The WM-tract-wise FC in the CN group were compared with every other group using unpaired-sample *t*-tests.

To measure the general trend of WM related functional connectivity as disease progresses, all the elements of each mFCM were averaged, providing a metric of overall-connectivity for each participant, and then the group mean and standard deviation of the overall-connectivity for each subject group were estimated and finally normalized by linear scaling.

The associations between each single FCM element and neuropsychological scores were evaluated by calculating the linear correlation coefficients between the element and each score across all participants. To gauge the overall correlation of a WM tract’s connectivity with the score, significant correlation coefficients along the same WM tract were summed.

To further evaluate the associations between combined FCM elements and each neuropsychological score, a random forest (RF) regression model was trained to predict the score after feature selection from all FCM elements. The value of goodness of fit, R^2^ was calculated based on comparing true and predicted scores.

### Machine Learning Classification

A support vector machine (SVM) with a radial-basis function kernel was used to classify the CN group and different combinations of groups of impaired subjects (i.e., ADD alone; LMCI and ADD together; MCI, LMCI and ADD together; EMCI, MCI, LMCI and ADD together; and SMC, EMCI, MCI, LMCI and ADD together). We used all FCM_WG_ and FCM_WW_ elements as initial features and implemented an RF algorithm to select those features that provided more accurate classifications [27]. In detail, the number of trees for the RF classifier was chosen to be 200 as it was observed that increasing the number of trees further resulted in no significant reduction of classification error. The splitting criterion for RF was based on the GUIDE algorithm [28]. Individual feature importance was computed by measuring how much the predictive accuracy of the RF classifier deteriorated when the feature’s values were randomly permuted ^2^. The idea is that altering the value of an important feature will degrade the performance of a classifier. After obtaining the importance of each feature individually, the features that did not improve performance at all were at first removed from the set. The remaining features were arranged in descending order of their importance. Features were added sequentially, and classification error was noted for this cumulative feature set. The optimal feature set was taken to be the one, which provided the lowest classification error. A similar method was used for feature selection in the case of regression analysis abovementioned. There were 5064 WM functional connectivities in this study, so the features, which were arranged in descending order of importance, were added five at a time sequentially to reduce the computational load. In the case of the classification task, SVM with a radial-basis function (RBF) kernel was optimized with respect to C and Gamma, the two hyper-parameters. C regularized the classifier, and Gamma denoted variance of the RBF kernel and controlled the width of the kernel. Mean squared error of a 10-fold cross-validation (CV) was calculated to measure the classification error. More specifically, the data were split into 10 subsets. The SVM model was trained on 9 subsets and then evaluated on the remaining subsets. This process was repeated 10 times, with a different subset as testing data each time. One error was estimated at each time and the final error was the average of the 10 errors. Also, the penalty involved for misclassification of the disease group versus control group was manually varied so that data imbalance between the groups did not tilt the model accuracy towards one group [29]. Moreover, the receiver-operating characteristic (ROC) analysis was performed and the area under curve (AUC), sensitivity, and specificity were noted.

Subjects that did not have any behavioral scores were removed from the regression study.

## Results

### Participant characteristics

**Table 2** shows characteristics of all 383 participants, grouped into CN (n=136), SMC (n=46), EMCI (n=83), MCI (n=37), LMCI (n=46) and ADD (n=35), in order of disease severity. Among these groups, no significant differences in age (P=0.93), sex (*P*=0.22), handedness (*P*=0.99), years of education (*P*=0.23), brain volume (*P*=0.41) or Hachinski scale (*P*=0.25) were observed. The scanner vendor breakdown, CSF amyloid beta, CSF Tau, CSF p-tau, FDG-PET, MMSE, CDR global, CDR-SOB, GDS, FAQ, WMS-LMII and ADAS-Cog scores did differ significantly among groups (*P* <0.007).

**Table 2.** Baseline Participant Characteristics.

| Characteristics | CN<br>(n=136) | SMC<br>(n=46) | EMCI<br>(n=83) | MCI<br>(n=37) | LMCI<br>(n=46) | ADD<br>(n=35) | P-value<br>(ANOVA or<br>chi-square) |
| --- | --- | --- | --- | --- | --- | --- | --- |
| Age, mean (SD), y | 74.5 (7.1) | 75.3 (5.8) | 74.5 (7.0) | 74.3 (7.2) | 74.6 (6.8) | 75.6 (6.8) | 0.93 |
| Female sex, No. (%) | 82(60) | 26(57) | 42 (51) | 15 (41) | 21 (46) | 16 (46) | 0.22 |
| Handedness, No. (%) | 126 (92) | 42 (91) | 76 (92) | 34 (92) | 42 (91) | 33 (94) | 0.99 |
| Education, mean (SD), y | 16.8 (2.3) | 16.8 (2.6) | 16.0 (2.7) | 16.3 (2.5) | 16.4 (2.9) | 16.2 (2.7) | 0.23 |
| Scanner vendor (S:G:P) | 86:27:24 | 18:15:13 | 31:11:41 | 18:9:10 | 19:6:21 | 6:3:26 | <0.001 |
| Brain volume (SD) X10<sup>5</sup> | 10.6 (0.9) | 10.7 (0.9) | 10.6 (2.2) | 10.4 (1.0) | 10.4 (1.1) | 10.2 (1.2) | 0.41 |
| CSF Amyloid beta (SD) X10<sup>2</sup> | 13.5 (4.3) | 13.8 (3.3) | 11.6 (4.6) | 11.4 (2.5) | 10.5 (4.0) | 6.7 (2.8) | <0.001 |
| CSF Tau | 261 (104) | 223 (78) | 273 (139) | 237 (113) | 275 (132) | 355 (130) | 0.007 |
| CSF p-Tau | 23.9 (10.6) | 19.9 (8.1) | 25.7 (15.5) | 23.2 (12.1) | 26.3 (14.3) | 34.6 (13.3) | 0.004 |
| FDG-PET | 1.34 (0.09) | 1.34 (0.09) | 1.30 (0.10) | 1.28 (0.13) | 1.26 (0.12) | 1.09 (0.14) | <0.001 |
| MMSE score, mean (SD) | 29.1 (1.7) | 29.1 (1.0) | 27.4 (2.9) | 28.0 (1.5) | 25.8 (5.6) | 22.4 (3.2) | <0.001 |
| CDR global score, mean (SD) | 0.0 (0.2) | 0.1 (0.2) | 0.4 (0.3) | 0.5 (0.0) | 0.6 (0.5) | 0.8 (0.2) | <0.001 |
| CDR-SOB score, mean (SD) | 0.3 (1.0) | 0.1 (0.4) | 1.7 (2.2) | 1.2 (0.8) | 2.6 (3.2) | 4.7 (1.5) | <0.001 |
| GDS score, mean (SD) | 0.8 (1.5) | 1.3 (1.3) | 1.8 (2.0) | 1.5 (1.3) | 1.8 (2.3) | 1.5 (1.3) | <0.001 |
| FAQ score, mean (SD) | 1.0 (3.7) | 0.4 (0.8) | 4.2 (6.5) | 3.1 (4.4) | 6.5 (8.7) | 14.6 (6.2) | <0.001 |
| WMS-LMII score, mean (SD) | 15.4 (3.3) | 15.4 (3.0) | 12.4 (5.1) | 9.2 (3.8) | 9.7 (5.0) | 4.5 (3.1) | <0.001 |
| ADAS-Cog score, mean (SD) | 9.6 (4.4) | 8.2 (3.1) | 12.0 (6.0) | 12.3 (3.4) | 14.0 (8.3) | 22.7 (7.4) | <0.001 |
| Hachinski scale, mean (SD) | 0.5 (0.7) | 0.6 (1.0) | 1.0 (1.2) | 0.8 (1.2) | 0.7 (0.9) | 0.9 (0.9) | 0.25 |

Note: S=Siemens; G=GE Medical Systems; P=Philips Medical System and Philips Healthcare. FDG-PET: average FDG-PET of angular, temporal, and posterior cingulate.

### WM functional connectivity at baseline

**Fig 1ac** shows the mean FCM_WG_ (mFCM_WG_) and mean FCM_WW_ (mFCM_WW_) for each clinical group. The overall patterns of mFCM_WG_ or mFCM_WW_ appear similar across the six groups. The mean correlation coefficient of each WM tract across GM ROIs or WM ROIs is illustrated in **Fig 1bd**, showing that cingulum (cingulate gyrus) (CGG), external capsule (EC) and internal capsule (IC) have higher correlations averaged over GM ROIs, and IC, splenium of corpus callosum (SCC), superior longitudinal fasciculus (SLF), and middle cerebellar peduncle (MCP) have higher WM-averaged correlations.

Comparisons between mFCM_WW_ (**Fig 1c**) and the group mean of GM-GM FCM (mFCM_GG_, **S2 Fig** in Supplement) shows a generally lower level of WM-WM correlation, with a relative decrease of 25-30% in overall average of FCM for every study group.

**Fig 1.**
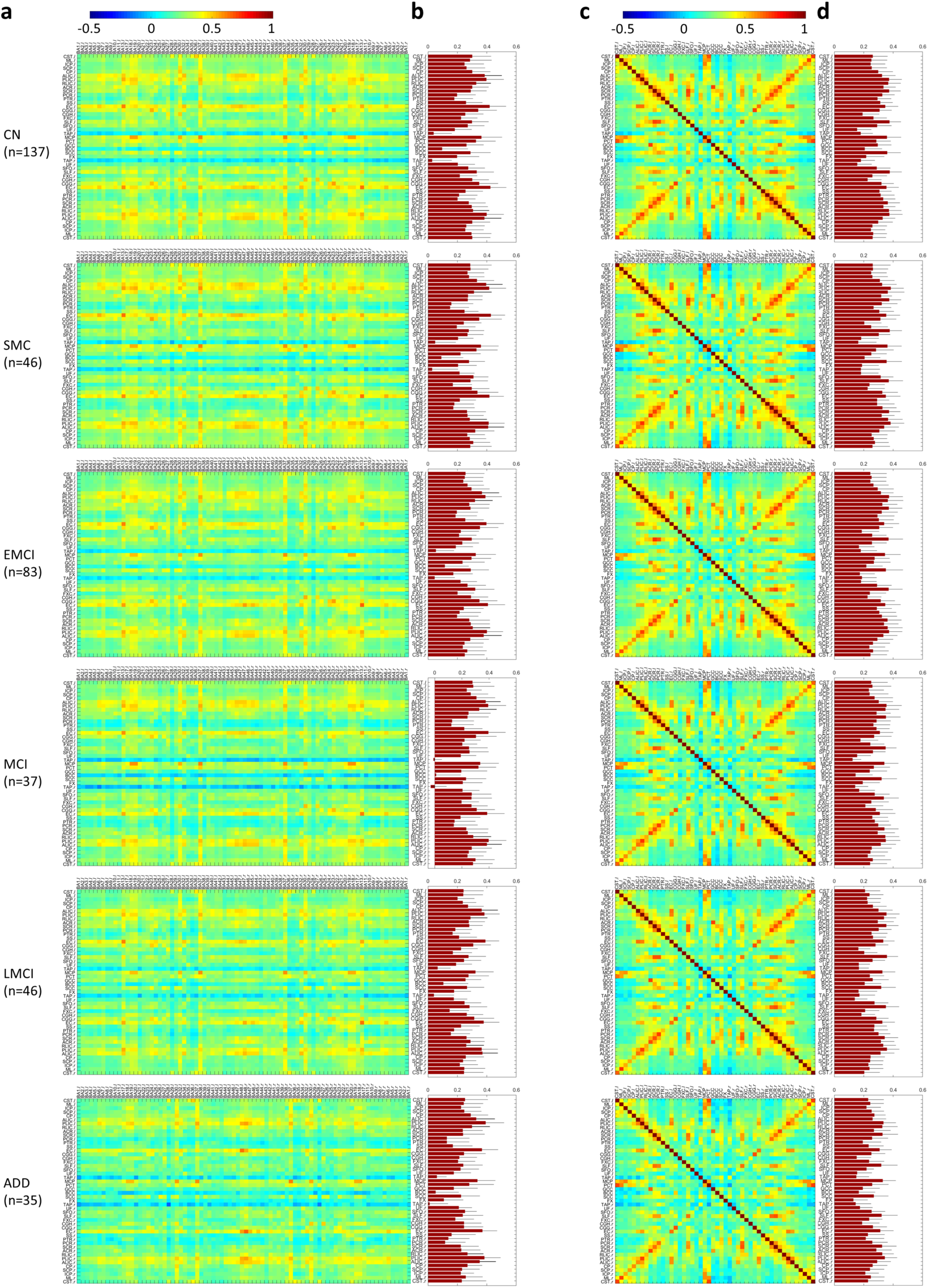
Mean of WM-GM and WM-WM functional correlation matrices (mFCM_WG_ and mFCM_WW_) as well as GM-averaged and WM-averaged correlation coefficients of WM tracts for each group were analyzed. Mean of FCM_WG_ (A) or FCM_WW_ (C) across all participants within group of CN, SMC, EMCI, MCI, LMCI or ADD. Each element in mFCM_WG_ (or mFCM_WW_) is the group mean correlation coefficient of the averaged BOLD time-courses between one WM region and one GM (or WM) region. GM-averaged (B) and WM-averaged (D) correlation coefficients of WM tracts for each group. Group mean (red bar) and standard deviation (black error bar) for each WM tract are shown. Full names of WM ROIs and GM ROIs are listed in **Table 1**.

### WM functional connectivity deficits in progression to AD

In LMCI or ADD patients, significant FC decreases were observed in a number of WM-GM pairs (**Fig 2a, f**) and WM-WM pairs (upper triangle in **Fig 2d, i**) compared with CN participants (*P_FDR_*<0.05) and there are obvious horizontal patterns in the difference matrices, corresponding to specific WM tracts. In particular, the corpus callosum (CC, including GCC, BCC and SCC), SLF*_lr_*, CGG*_lr_*, sagittal stratum (SS*_lr_*), posterior thalamic radiation (PTR*_lr_*) and corona radiata (CR*_lr_*, including ACR, SCR and PCR) show profound declines in WM-GM connectivity in the ADD groups relative to the CN group (**Fig 2g**), consistent with previous findings showing microstructural degeneration detected by diffusion MRI in CC, SLF, CGG, SS, PTR and CR in ADD patients[30]. The SS*_lr_*, fornix (cres) (FXC*_lr_*), SLF*_lr_*, right hippocampal cingulum (CGH*_r_*), EC*_r_*, PTR*_r_* and PCR*_r_* show significant decreases in WM-GM connectivity in the LMCI group relative to CN group (**Fig 2b**). The CC, SLF*_lr_*, CGH*_lr_*, CGG*_lr_*, EC*_r_*, SS*_lr_*, PTR*_lr_*, PCR*_lr_*, SCR*_lr_*, ACR*_l_*, RLIC*_lr_* and PLIC*_lr_* show significant declines in inter-tract FC in both the LMCI and ADD groups relative to the CN group (**Fig 2e, j**). Moreover, the effect sizes of group deficits with *P*_FDR_<0.05 were mostly larger than 0.3 (**Fig 2c, h** and lower triangle in **Fig 2d, i**).

**Fig 2.**
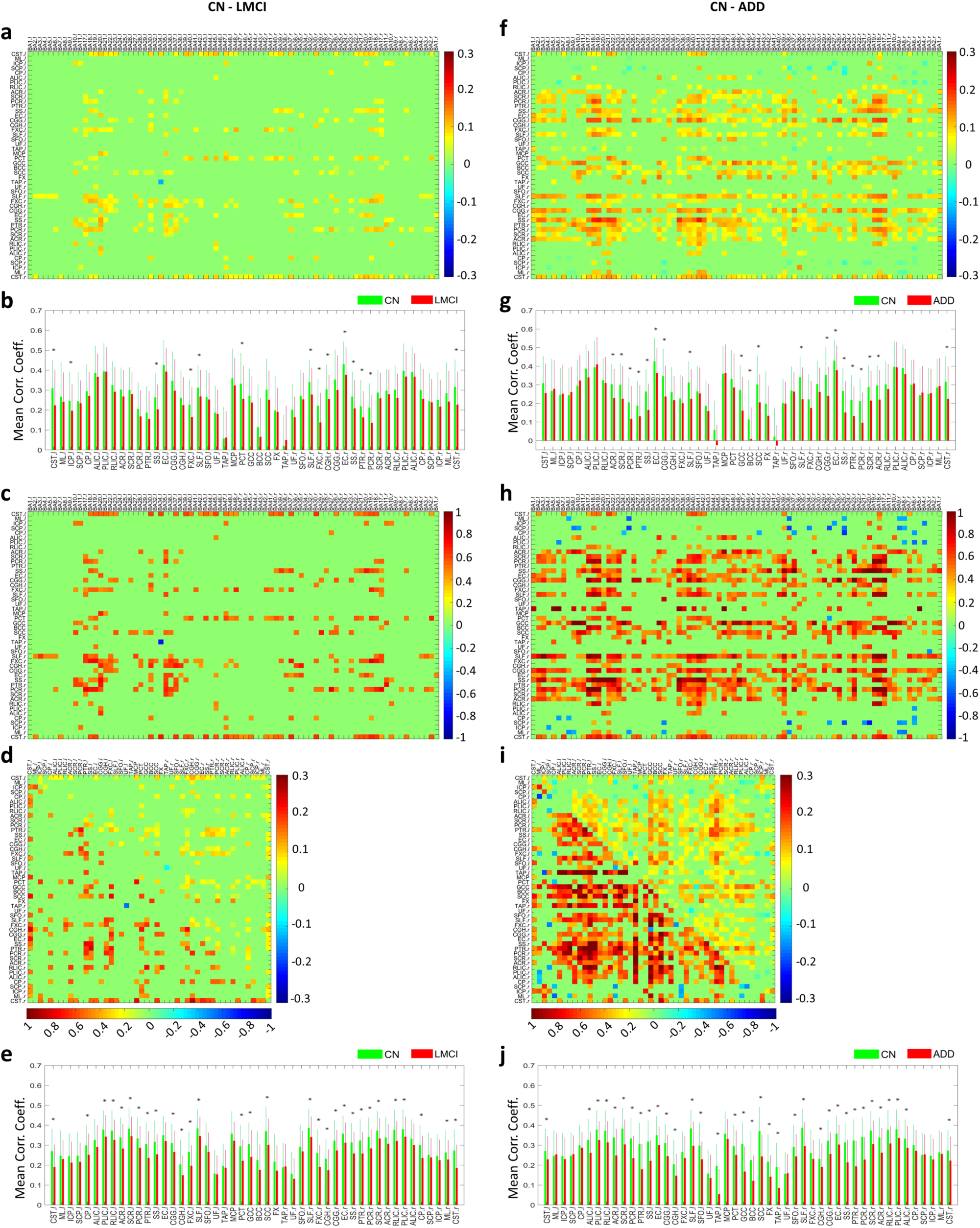
Significant differences of mFCM_WG_ and mFCM_WW_ between controls and impaired groups. (**a, f**) Difference of subtracting mFCM_WG_ of LMCI (**a**) or ADD (**f**) from mFCM_WG_ of CN group. The *P*-value for each element was derived from permutation-test (10,000 permutations) across all participants within groups, and then adjusted using an FDR. Those elements with *P_FDR_* >0.05 were set to be zero. (**b, g**) GM-averaged correlation coefficients of WM tracts in CN group (green) and LMCI group (red) (**b**) and in CN group (green) and ADD group (red) (**g**). Mean and standard deviation for each WM tract are shown, and black asterisks (*) denote *p* <0.05 calculated by unpaired-sample *t-*test of mean correlation coefficients. (**c, h**) Effect size of the mFCM_WG_ difference between CN and LMCI (**c**) or ADD (**h**), thresholded by *P_FDR_*. (**d, i**) Difference of mFCM_WW_ between CN and LMCI or ADD (upper triangle) and effect size of the mFCM_WW_ difference (lower triangle). (**e, j**) WM-averaged correlation coefficients of WM tracts in CN group (green) and LMCI group (red) (**e**) and in CN group (green) and ADD group (red) (**g**).

By contrast, no significant changes in FCM_WG_ or FCM_WW_ between CN and any of the early disease groups (i.e., SMC, EMCI and MCI) were observed at the same *P*-value threshold. But their mean *P_FDR_* were 0.99, 0.65, and 0.22, respectively, suggesting a progression towards significant differences. By contrast, previous studies on brain microstructure reported alterations in a few selected WM regions in MCI participants, involving cingulate bundle, inferior FOF and parahippocampal subgyral fibers [31, 32], where connectivity deficits were found in our LMCI and ADD cases.

### Correlation between WM functional connectivity and neuropsychological scores

The normalized group means of overall-connectivity decreased gradually until the MCI stage, and then fell rapidly to very low values as MCI progressed to LMCI and ADD stages (**Fig 3a**), a behavior which conforms to current hypothetical models of AD evolution [33]. The corresponding normalized overall trends in 7 behavioral measures across groups (**Fig 3b**) show a striking similarity with this overall-connectivity trend.

**Fig 3.**
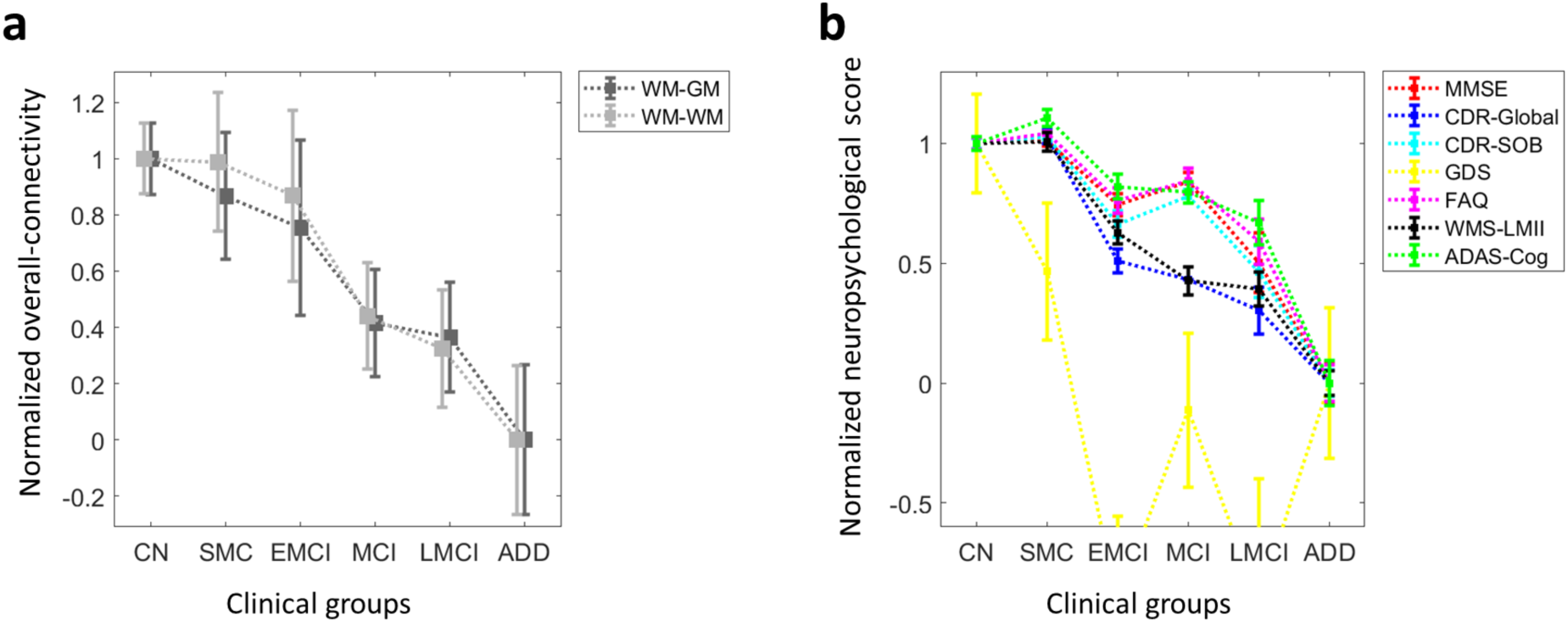
Normalized overall-connectivity and neuropsychological scores for each clinical group in AD progression. (**a**) Normalized group mean (gray square) and standard deviation of mean (gray bar) of the overall-connectivity for each clinical group. The six clinical groups are CN, SMC, EMCI, MCI, LMCI and ADD groups. (**b**) Normalized mean (colored square) and standard deviation (colored bar) of the neuropsychological scores for each clinical group.

**Fig 4a-d** shows correlation coefficients between elements in FCM_WW_ or FCM_WG_ and neuropsychological scores which are significantly different from zero (*P_FDR_*<0.05). No significant correlations were found between any element in FCM and the Hachinski scale or GDS score.

**Fig 4.**
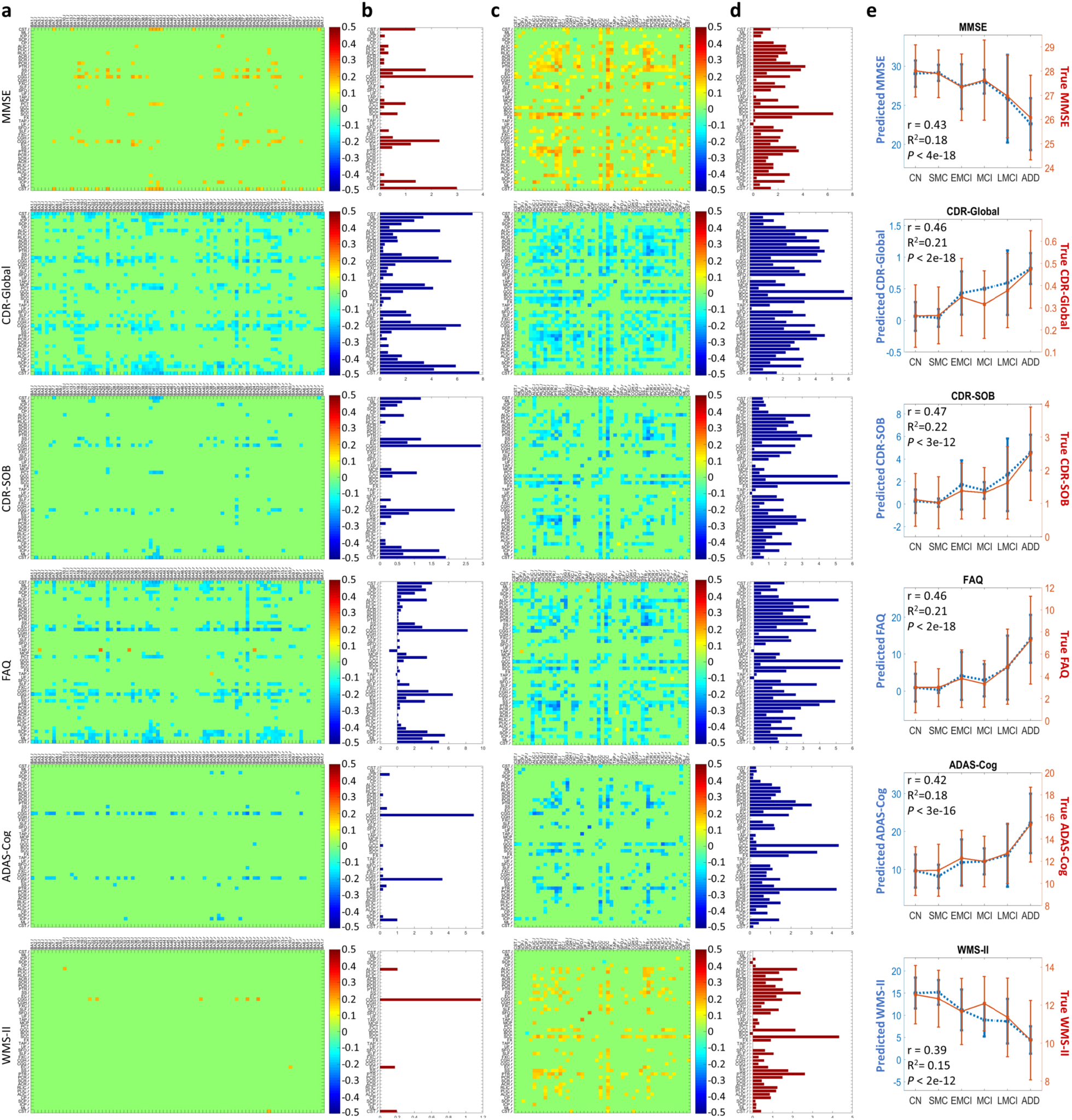
Correlations between WM functional connectivity and neuropsychological scores across all subjects. (**a or c**) Matrix of Pearson’s correlation coefficients between single elements in FCM_WG_ or FCM_WW_ and MMSE score, CDR-Global score, CDR-SOB score, FAQ score, ADAS-Cog score, or WMS-LMII score. Each correlation coefficient with *P_FDR_* > 0.05 was set to be zero. (**b** or **d**) Sum of significant correlation coefficients along each WM tract in **a** or **c**. See **Table 1** for the lists of WM and GM ROIs. (**e**) Group means and standard deviations of true scores and predicted scores using RF regression model with all WM functional connectivity as initial features. The r, R^2^ and *P* in each plot are the Pearson’s correlation coefficient between true scores and predicted scores across all subjects, R-square value and *P*-value, respectively.

FCM_WG_ and FCM_WW_ elements within SS*_lr_*, CGG*_lr_*, CGH*_l_*, FXC*_lr_*, SCC, PTR*_lr_*, and bilateral cerebral spinal tract (CST*_lr_*) showed significant positive associations with MMSE scores [34] (**Fig 4a-d**), indicating reduced WM FC corresponds to more severe cognitive impairment. A previous ADNI study found that MMSE scores were strongly associated with a DTI index [35] in SS*_lr_*, CGG*_l_*, CGH*_lr_*, FXC, and SCC. Another study demonstrated WM micro-structural damage was correlated with MMSE, most strongly in temporal lobe [36] where the CGH, FXC and part of SS reside. On the other hand, the lower WM functional connectivity might be also due to damage to GM regions to which the WM tracts connect, including cellular pathology [4] and gross atrophy [37]. GM loss was reported to be strongly correlated with decreases in MMSE in all regions that showed prominent GM atrophy in AD [38].

The CDR-global [39] and CDR-SOB scores are indicators of dementia severity [40]. The sum of negative correlations along WM tracts between FCM and CDR-global was strong in SS*_lr_*, CGG*_lr_*, FXC*_lr_*, SLF*_r_*, SCC, GCC, FX, PCR*_lr_*, PTR*_lr_*, ALIC*_l_*, and CST*_lr_* (**Fig 4a-d**). For the linkage between FCM elements and CDR-SOB score, clear negative correlations were found in SS*_lr_*, CGG*_lr_*, FXC*_l_*, PTR*_lr_*, PCR*_lr_*, GCC, SCC, ALIC*_l_*, RLIC*_l_*, and CST*_r_*. A previous study found that diffusivity measures were strongly associated with CDR-SOB in SS*_lr_*, CCG*_l_*, FXC*_l_*, PTR*_l_*, PCR*_l_* and the entire CC [35].

Negative correlations were dominant in SS_lr_, CGG_lr_, PTR*_lr_*, PCR*_lr_*, SLF*_r_*, ALIC*_l_*, PLIC*_l_*, GCC and SCC between FCM and FAQ [41] that describe the level of performance of daily function activities (**Fig 4a-d**).

CGG*_lr_* showed the most significant negative correlations between FCM_WG_ and the ADAS-Cog [42] score, the overall degree of cognitive decline (**Fig 4a-d**). GCC, SCC, FX, PTR*_lr_*, PCR*_l_*, CGG*_l_*, SS*_l_*, showed significant correlation between FCM_WW_ and ADAS-Cog (**Fig 4cd**). A previous study found that diffusivity measures were strongly correlated with ADAS-Cog in GCC, SCC, FXC*_l_*, PTR*_lr_*, CGG*_l_*, and SS*_lr_*, among others [35].

SCC, GCC, SS*_lr_*, ALIC*_l_*, PTR*_r_*, and CGG*_l_* showed stronger positive correlations between FC and the WMS-LMII [43] score, a measure of episodic memory (**Fig 4a-d**). CGG and CGH constitute the major pathways between the hippocampal regions and posterior cingulate cortex (PPC). The intrinsic FC between hippocampus and PPC has been reported to be closely associated with WMS-LM scores in elderly people [44]. A portion of SS connects to temporal lobe and prefrontal cortex which are also engaged in episodic memory processing [45]. Moreover, WM tracts in left hemisphere appear to correlate with WMS-LMII scores more widely and strongly (**Fig 4a-d**), consistent with previous findings that verbal memory tasks are more sensitive to left hemisphere dysfunction [46] and damage to left temporal lobe has consistently been associated with an impairment of verbal memory [47].

### Correlation between combined WM functional connectivities and neuropsychological scores

The correlation coefficients between the true and RF-predicted scores of ADAS-Cog, CDR-Global, CDR-SOB, FAQ and MMSE were 0.39-0.47 with highly significant *P*-value (<0.001) (**Fig 4e**). The R^2^ values in **Fig 4e** indicate that 15%-22% of the variances of those individual scores could be explained by the variance of the overall combined WM functional connectivities, and vice versa.

### Prediction of AD stages

The performance of the SVM classification using WM FC features was best for distinguishing ADD from CN group with sensitivity of 0.83 and specificity of 0.81 (AUC=0.87). The performance using optimized features reduced monotonically with addition of patients from earlier stages to the ADD group (**Fig 5**). Also, feature selection helped to improve performance in all the classifications.

**Fig 5.**
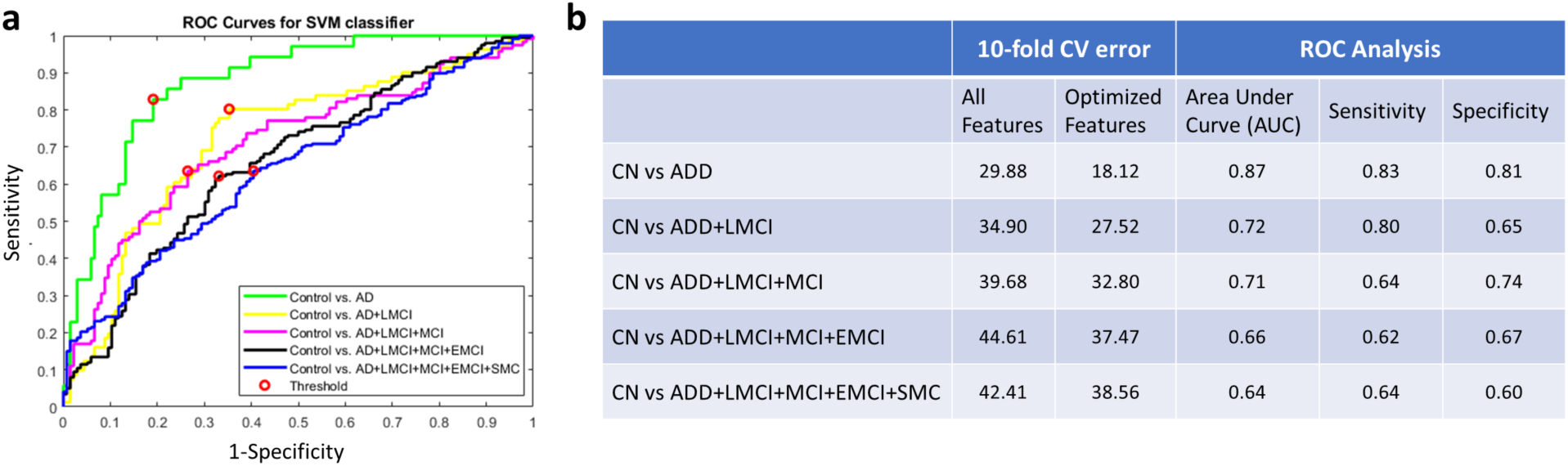
ROC curves of SVM classifications and a summary of their CV errors, AUC, sensitivity and specificity. (**a**) ROC curves of SVM algorithm for distinguishing patients from CN. Difference color represents different cumulative group of patients. (b) The errors of 10-fold CV and ROC related indices-AUC, sensitivity and specificity for the classifications.

## Discussion

In conventional rsfMRI studies, correlations in BOLD signals between GM regions are interpreted as revealing FC. Extending this concept, we investigated the FC between WM and GM regions or between WM regions for 383 ADNI participants from six subject groups. We mainly found that: 1) LMCI and ADD have significant deficits in regional WM FC relative to CN, 2) regional WM FC is significantly associated with neuropsychological scores (i.e., MMSE, CDR, FAQ, ADAS-Cog and WMS-LMII), and 3) WM FC can be used to distinguish between patients and controls. To the best of our knowledge, this is the first investigation of novel measures of functional connectivity and degeneration in WM throughout the evolution of AD pathology. Our findings indicate that FC of WM from MRI may serve as a novel in vivo biomarker to identify changes in brain in AD.

In our analyses, we constrained ROIs to GM or WM only in order to avoid partial volume averaging effects which potentially could overestimate the correlation of time-courses between regions. These correlations of WM with GM or other WM volumes are unlikely caused by drainage effects from adjacent GM because cortical drainage occurs outwards, towards the brain surface while deeper WM veins drain inwards to sub-ependymal veins near the lateral ventricles, so there is no direct vascular communication between them [48]. In our analyses we did not regress out global signals because there is growing evidence that they may contain valuable information [49, 50]. Other physiological noises (such as that caused by variations in heart rate and respiration) were, however, regressed out. With these factors in mind, we believe that the WM FC we measured is neither noise effect nor simply a reflection of GM changes.

From a pathophysiological perspective, the decreases in WM FC metrics observed in late AD stages may be directly attributable to GM abnormalities, WM degeneration, metabolic changes and/or cerebrovascular changes. GM abnormalities in AD include the appearance of neurofibrillary/senile plaques, neuronal loss, cell shrinkage, reduced dendritic density and synaptic losses [51, 52]. The consequent neural dysfunction could lead to less engagement of WM in transmission of neural information. WM degeneration during AD progression [53-55] includes demyelination [56] and axonal damage [57], which may also weaken the ability of WM to transfer neural signals between regions. WM degeneration has been reported as a direct consequence of amyloid deposition and tau phosphorylation in GM, and of damage to oligodendrocytes, possibly initiated by ischemia, excitotoxicity, oxidative stress and/or iron overload [58]. Cerebral hypometabolism is found in MCI and AD patients throughout limbic structures, involving hippocampal complex, medial thalamus, mammillary bodies and posterior cingulate. AD patients may also have hypometabolism in amygdala, temporoparietal and frontal association cortex [59, 60]. Cerebral hypoperfusion defects in AD are severe [61], and atherosclerosis that leads to cerebral hypoperfusion is significantly correlated with AD severity. Both hypometabolism and hypoperfusion may have direct effects on BOLD signals in GM and WM. Further studies to understand the relationship between the GM changes and decreases in WM connectivity in AD are clearly needed and may reflect a combination of factors including direct causes and effects, comorbidities and mutual dependences.

The performances of our SVM classifications indicate that it is feasible to differentiate between CN and AD patients objectively using WM connectivity metrics only. The performance decreases as patients at earlier stages of AD are included sequentially, because there are smaller reductions in WM connectivity early in the disease. The decrease in classification errors using an optimal feature set emphasizes the importance of removing redundant features in the analysis i.e. those WM connectivities that do not contribute new information for the classification. It will be interesting to study in future those specific WM tracts that provide complementary information for maximal differentiation between groups.

## Conclusions

The present study indicates that WM functional connectivities 1) decline regionally in LMCI and ADD groups relative to a CN group, 2) are significantly related to cognitive scores, and 3) can serve as machine learning features for distinguishing between AD patients and CN with an acceptable sensitivity and specificity. These findings suggest the potential of WM FC, which has been largely overlooked to date, as a novel neuroimaging biomarker to assess AD progression.

## Data Availability

The analyzed data may be available upon request. Please contact Dr. Gao for further information.

## Acknowledgments

The project is supported by the NIH grant R01 NS093669 and a Vanderbilt University Discovery Grant 600670. We also thank the Vanderbilt Advanced Computing Center for Research and Education (ACCRE) for the support of cluster computation.

Data collection and sharing for this project was funded by the ADNI (NIH Grant U01 AG024904) and DOD ADNI (DOD award number W81XWH-12-2-0012). ADNI is funded by the National Institute on Aging, the National Institute of Biomedical Imaging and Bioengineering, and through generous contributions from the following: AbbVie, Alzheimer’s Association; Alzheimer’s Drug Discovery Foundation; Araclon Biotech; BioClinica, Inc.; Biogen; Bristol-Myers Squibb Company; CereSpir, Inc.; Cogstate; Eisai Inc.; Elan Pharmaceuticals, Inc.; Eli Lilly and Company; EuroImmun; F. Hoffmann-La Roche Ltd and its affiliated company Genentech, Inc.; Fujirebio; GE Healthcare; IXICO Ltd.; Janssen Alzheimer Immunotherapy Research & Development, LLC.; Johnson & Johnson Pharmaceutical Research & Development LLC.; Lumosity; Lundbeck; Merck & Co., Inc.; Meso

Scale Diagnostics, LLC.; NeuroRx Research; Neurotrack Technologies; Novartis Pharmaceuticals Corporation; Pfizer Inc.; Piramal Imaging; Servier; Takeda Pharmaceutical Company; and Transition Therapeutics. The Canadian Institutes of Health Research is providing funds to support ADNI clinical sites in Canada. Private sector contributions are facilitated by the Foundation for the NIH (www.fnih.org). The grantee organization is the Northern California Institute for Research and Education, and the study is coordinated by the Alzheimer’s Therapeutic Research Institute at the University of Southern California. ADNI data are disseminated by the Laboratory for Neuro Imaging at the University of Southern California.

